# The size of myocardial infarction and peri-infarction edema are not major determinants of diastolic impairment after acute myocardial infarction

**DOI:** 10.1101/2022.07.01.22277049

**Authors:** Martin G Sundqvist, Dinos Verouhis, Peder Sörensson, Loghman Henareh, Jonas Persson, Nawzad Saleh, Magnus Settergren, Nils Witt, Felix Böhm, John Pernow, Per Tornvall, Martin Ugander

## Abstract

**Aims:** Diastolic dysfunction after myocardial infarction (MI) is a marker of poor prognosis. The relationship between myocardial infarction size (IS), myocardial edema, and diastolic dysfunction is poorly understood, both in the acute phase, and in the development of diastolic dysfunction in the follow-up setting. Using a mechanistic approach could potentially add insights.

**Methods and results:** Patients underwent cardiovascular magnetic resonance (CMR) imaging and echocardiography including mechanistic analysis using the parameterized diastolic filling method within 4-7 days (acute) and 6 months after a first acute anterior MI (n=74). Linear regression modeling of echocardiographic diastolic parameters using CMR IS with and without inclusion of the myocardium at risk (MAR) and model comparisons with likelihood ratio tests were performed. Diastolic parameters at 6 months follow-up were modelled using final IS. For most parameters there was no association with acute IS, except for deceleration time (R^2^=0.24, p<0.001), left atrial volume index (R^2^=0.13, p=0.01) and the mechanistic stiffness parameter (R^2^=0.21, p<0.001). Adding MAR improved only the e′ model (adjusted R^2^ increase: 0.08, p=0.02). At 6 months follow-up, final IS was only associated with viscoelastic energy loss (R^2^=0.22, p=0.001).

**Conclusion:** In acute MI, both IS and MAR are related to diastolic function but only to a limited extent. At 6 months after infarction, increasing IS is related to less viscoelastic energy loss, albeit also to a limited extent. The relationship between IS and diastolic dysfunction seems to be mediated by mechanisms beyond simply the spatial extent of ischemia or infarction.

## Introduction

It is established that severe diastolic dysfunction, in the form of a restrictive filling pattern, is a marker of poor prognosis following myocardial infarction (MI)^1^. Furthermore, infarct size (IS) is a major determinant of prognosis^2–4^. Intuitively, one could assume that larger MIs would be associated with worse diastolic function. Previous research in this area has, however, indicated that the relationship between acute or near-term follow-up IS and the parameters used to assess diastolic function is weak^5,6^. One possible factor that could influence this relationship is the myocardial edema that surrounds the infarcted tissue in the acute phase, which corresponds to the myocardial area at risk (MAR), as it has been shown that myocardial edema can impair systolic function and cause increased myocardial stiffness^7,8^. To our knowledge, only two previous studies have investigated the relationship between the extent of myocardial edema and left ventricular diastolic function after MI. One study used continuous echocardiographic measures of diastolic function and found no association^9^, whereas the other found an association, albeit using diastolic dysfunction classified into groups^10^, thus limiting the potential to elucidate pathophysiological mechanistic relationships. Furthermore, while previous studies demonstrated that ongoing myocardial ischemia affected diastolic function assessed as invasively measured increased ventricular stiffness^11^, the time course of how a manifest MI impacts the development of diastolic dysfunction is not known. It is also plausible that the relationship between IS and diastolic function could be obscured in the acute setting, as loading conditions, medication, and time elapsed since ischemia can vary considerably among patients. An assessment in a more stable phase could possibly yield more accurate results, as well as an opportunity to examine the change over time.

In the present study, we aimed to examine to what extent IS and MAR measured by cardiovascular magnetic resonance (CMR) imaging influenced various echocardiographic parameters used to assess diastolic function. This was evaluated both using conventional echocardiographic measures, and also by using a more mechanistic approach by employing the parameterized diastolic filling (PDF) method^12^.

## Methods

### Patients

The patients in this study were included in the RECOND trial^13^, which examined the effect of remote ischemic conditioning in patients presenting with a first anterior ST-elevation myocardial infarction (STEMI). The RECOND trial did not find evidence for any effect of the investigated treatment on IS or myocardial salvage, and included patients over 18 years of age presenting with anterior STEMI leading to percutaneous coronary intervention (PCI). Exclusion criteria included prior MI or coronary artery bypass grafting, left bundle branch block, atrial fibrillation, and contraindications to CMR imaging. The patients were examined with CMR at 4-7 days and at 6 months after MI. Echocardiography was performed within 24 hours of the first CMR examination, and again within 2 weeks of the second CMR study.

### CMR

CMR was performed using a 1.5T scanner (Magnetom Aera, Siemens Healthcare, Erlangen, Germany). A gadolinium-based contrast agent (gadopentetate dimeglumine, 0.2 mmol/kg, Guerbet, France) was administered intravenously. Early contrast-enhanced steady-state free precession and late gadolinium enhancement (LGE) images were obtained in contiguous short axis views (8 mm thickness, 2 mm gap) covering the entire left ventricle, and in the 2-, 3-, and 4-chamber long axis views, for the determination of MAR and IS, respectively. LGE images were acquired 15 to 20 minutes after contrast injection using a phase sensitive inversion recovery gradient echo sequence. CMR image analyses were performed off-line using the freely available software Segment v1.9 R3967 (http://segment.heiberg.se)^14^. MAR was manually outlined in both end-diastolic and end-systolic phase using early contrast-enhanced steady state free precession images^15^. The same short-axis stack was used for calculating left ventricular volumes, ejection fraction, stroke volume, and mass. Epi- and endocardial borders were manually outlined on LGE images. IS was quantified using the automated Otsu quantification method^16^, with weighted approach^17^, followed by manual adjustments if needed. CMR examinations were interpreted by two observers blinded to the randomization of the RECOND trial.

### Echocardiography

Echocardiography was performed using Vivid E9 (General Electric Healthcare, Horten, Norway) scanners. All examinations were analyzed in EchoPAC v1.13 (General Electric Healthcare, Horten, Norway). Registrations were performed in accordance with clinical guidelines. For PDF analysis, extended registrations of transmitral flow during free breathing were recorded. PDF analysis was performed using the freely available software Echo E-waves v1.0 (http://www.echoewaves.org)^18^. All echocardiographic analyses were performed by one observer (MGS).

### The PDF method

The PDF method models early diastolic LV filling as a case of damped harmonic motion, and by using the E-wave obtained from pulsed wave (PW) Doppler registration of early diastolic mitral inflow as input, parameters reflecting LV diastolic stiffness (*k*), viscoelastic energy loss (*c*), and load (*x*_0_) can be obtained^12^. Briefly, the LV is compressed during systole, after which, defining the beginning of the early diastolic filling phase, the LV begins to recoil. The recoil leads to a fall in LV pressure which, as LV pressure drops below left atrial pressure, sucks blood into the LV, constituting the early filling phase of diastole. Using PW Doppler, the velocity of blood entering the LV is registered. The E-wave thus describes the speed of early LV filling, and by curve fitting the PDF formula (see Supplemental material) to the E-wave contour, three parameters (*x*_0_, *k*, and *c*) can be calculated. In the PDF framework, the amount of compression, or load, is denoted *x*_0_, and the product of *x*_0_ and the stiffness parameter *k* (*kx*_0_) gives the initial force driving recoil (peak driving force), and the energy of the system is given by 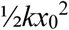. The recoiling motion will be dampened by various factors, mainly the energy loss associated with impaired relaxation and viscoelastic properties of the LV, lumped in the damping parameter *c*. At the time of maximal E-wave velocity, the force resisting recoil reaches its peak (peak resistive force), and is given by the product of *c* and the maximal velocity of the E-wave (*c*V_max_) The influence of the energy loss can also be expressed as a kinematic filling efficiency index (KFEI) which is ratio of the actual E-wave velocity time integral (VTI) to the theoretical E-wave VTI which would have resulted with no energy loss during recoil (*c*=0) ^19^, or as an approximation of tau ^20^, which is linearly correlated to the prolongation of the deceleration time that is caused by the energy loss represented by *c*.

### Statistical analysis

Patient characteristics including CMR data and echocardiographic diastolic parameters are presented as median [interquartile range] for continuous data, and as counts and percentages for categorical variables. Regression models investigating the relationship between acute IS, MAR, and the diastolic parameters were fitted with a pre-specified complexity comprising three-knot restricted cubic splines in order to accommodate eventual non-linear relationships^21^. For each diastolic parameter as a dependent variable, two models were fitted: one with only acute IS as an independent variable, and one with acute IS and MAR as independent variables. To test whether the addition of MAR improved the goodness of fit, a likelihood ratio test was used on each pair of models. Similarly, follow-up diastolic parameters were modelled with 6-month IS as the independent variable. The difference in unadjusted R^2^ between the IS only and IS+MAR models was also demonstrated graphically. All statistical analyses were performed in R v4.1.3 (R Core Team, 2022)^22^, regression modeling was done using the rms package^23^, and figures produced using the ggplot2 package^24^. A p-value <0.05 was interpreted as being statistically significant.

## Results

Ninety-three patients were included in the RECOND trial based on having a CMR examination of sufficient quality^13^. Of these, 74 patients had an echocardiogram performed within 1 [0–1] day from the first examination, and 61 patients had a follow-up echocardiogram at 6 months. Patient baseline characteristics and CMR results are presented in Table 1. The difference between MAR and acute IS was on average 18.7±7.0 percentage points of LV mass. The results from analysis of the diastolic parameters at baseline and at follow-up are presented in Table 2. As a registration of the tricuspid regurgitation velocity was possible in only 22% of patients, this parameter was not used in further analyses. A typical example of the application of the PDF method is shown in Figure 1. The relationships between IS and the diastolic parameters in the acute setting and at follow-up are illustrated in Figure 2A for the conventional parameters and in Figure 2B for the PDF parameters. The results from regression modeling of the relationship between the diastolic parameters and acute IS and MAR are presented in Table 3. The differences in unadjusted R^2^ between the model pairs are illustrated in Figure 3. The results from modeling of the diastolic parameters at follow-up are presented in Table 4.

**Table 1.** Baseline and CMR data.

|  |  |
| --- | --- |
| Number of subjects, n | 74 |
| Age, years | 61 [55–68] |
| Male sex, n (%) | 70 (95) |
| Hypertension, n (%) | 24 (32) |
| Diabetes, n (%) | 8 (11) |
| Infarction size, % of LV | 17.9 [13.5–25.7] |
| Myocardium at risk, % of LV | 42.0 [33.3–47.8] |
| LVEDV, ml | 182 [160–219] |
| LV ejection fraction, % | 50 [44–54] |

Data are presented as median [interquartile range] or n (%). LV – left ventricle. LVEDV – left ventricular end-diastolic volume

**Table 2.** Diastolic parameters from echocardiography at baseline and follow-up.

| Parameter | Baseline | Follow-up |
| --- | --- | --- |
| Number of subjects | 74 | 61 |
| E/A | 1.26 [1.05–1.62] | 1.30 [0.97–1.49] |
| e', cm/s | 7.6 [6.2–8.5] | 8.2 [6.8–9.6] |
| E/e' | 9.9 [8.4–12.1] | 8.5 [7.5–10.6] |
| DT, ms | 169 [150–191] | 193 [176–210] |
| LAVI, ml/m<sup>2</sup> | 31 [27–35] | 33 [28–40] |
| TR Vmax, m/s | 2.5 [2.2–2.9] | 2.5 [2.2–2.6] |
| *Diastolic dysfunction grading* |  |  |
| Normal, n (%) | 26 (35) | 36 (59) |
| Indeterminate, n (%) | 11 (15) | 12 (20) |
| Diastolic dysfunction, n (%) | 37 (50) | 13 (21) |
| Increased LVP, n (%) | 8 (11) | 3 (5) |
| *PDF parameters* |  |  |
| c, g/s | 17.2 [14.8–19.7] | 16.9 [15.0–18.6] |
| k, g/s<sup>2</sup> | 213 [171–254] | 166 [138–198] |
| x<sub>0</sub>, cm | 9.4 [8.0–11.1] | 11.4 [10.3–12.8] |
| KFEI, % | 54.6 [53.3–56.4] | 53 [52–55] |
| Tau, ms | 61 [57–67] | 71 [64–77] |
| Energy, mJ | 0.94 [0.68–1.26] | 1.14 [0.86–1.35] |
| Peak driving force, mN | 20.2 [15.7–24.3] | 19.6 [17.1–22.7] |
| Peak resistive force, mN | 11.6 [9.0–14.7] | 11.8 [10.5–14.5] |

Data presented as median [interquartile range] or n (%). DT – deceleration time. LAVI – left atrial volume index. TR Vmax – tricuspid regurgitation maximal velocity. LVP – left ventricular filling pressure. KFEI – kinematic filling efficiency index.

**Table 3.** Regression model results, acute setting (n=74).

|  | Models with only IS as predictor |  |  | Models with IS and MAR as predictors |  |  |  | Model comparisons |  |
| --- | --- | --- | --- | --- | --- | --- | --- | --- | --- |
| Parameter | IS effect | R<sup>2</sup> | p-value* | IS effect | MAR effect | R<sup>2</sup> | p-value* | Adjusted R<sup>2</sup> difference | Likelihood ratio test p-value |
| E/A | 0.08<br>[-0.12, 0.28] | 0.01 | 0.73 | 0.04<br>[-0.28, 0.35] | 0.09<br>[-0.25, 0.44] | 0.01 | 0.92 | -0.02 | 0.83 |
| e', cm/s | -0.5<br>[-1.1, 0.1] | 0.04 | 0.28 | 0.4<br>[-0.5, 1.3] | -0.8<br>[-1.8, 0.2] | 0.14 | **0.04** | 0.08 | **0.02** |
| E/e' | 0.5<br>[-0.9, 1.9] | 0.01 | 0.72 | -0.3<br>[-2.4, 1.7] | 0.3<br>[-1.9, 2.5] | 0.05 | 0.48 | 0.01 | 0.22 |
| LAVI, ml/m<sup>2</sup> | -1<br>[-4, 1] | 0.13 | **0.01** | -2<br>[-6, 2] | 3<br>[-1, 7] | 0.17 | **0.03** | <0.01 | 0.30 |
| DT, ms | -26<br>[-38, -14] | 0.24 | **<0.001** | -27<br>[-46, -8] | 7<br>[-14, 27] | 0.25 | **0.001** | -0.01 | 0.60 |
| c, g/s | 0.7<br>[-0.6, 2.0] | 0.06 | 0.13 | -1.0<br>[-3.0, 1.1] | 2.3<br>[0.0, 4.5] | 0.12 | 0.07 | 0.04 | 0.08 |
| k, g/s<sup>2</sup> | 35<br>[14, 55] | 0.21 | **<0.001** | 25<br>[-7, 56] | 20<br>[-15, 55] | 0.22 | **0.002** | -0.01 | 0.49 |
| x<sub>0</sub>, cm | -0.9<br>[-1.8, 0.04] | 0.05 | 0.16 | -0.8<br>[-2.2, 0.7] | -0.5<br>[-2.2, 1.1] | 0.06 | 0.36 | -0.02 | 0.68 |
| KFEI, % | 1.0<br>[-0.2, 2.3] | 0.04 | 0.22 | 2.2<br>[0.2, 4.1] | -1.5<br>[-3.7, 0.6] | 0.07 | 0.25 | 0.01 | 0.27 |
| Tau, ms | -5<br>[-9, -1] | 0.08 | 0.05 | -10<br>[-16, -4] | 5<br>[-1, 12] | 0.13 | **0.04** | 0.03 | 0.11 |
| Energy, mJ | 0.00<br>[-0.18, 0.17] | <0.01 | 0.99 | -0.06<br>[-0.33, 0.22] | 0.01<br>[-0.30, 0.31] | 0.01 | 0.94 | -0.02 | 0.67 |
| Peak driving force, mN | 1.6<br>[-0.5, 3.7] | 0.05 | 0.16 | 1.1<br>[-2.2, 4.3] | 0.2<br>[-3.4, 3.8] | 0.06 | 0.40 | -0.02 | 0.80 |
| Peak resistive force, mN | 0.7<br>[-0.7, 2.1] | 0.03 | 0.32 | -0.1<br>[-2.2, 2.0] | 0.7<br>[-1.7, 3.0] | 0.05 | 0.50 | -0.01 | 0.55 |

Effects are the predicted differences in dependent parameters comparing the 1<sup>st</sup> and 3<sup>rd</sup> quartile of IS and MAR, with 95% confidence intervals.
\*) These p-values stem from the F-test of overall significance for the models. LAVI – left atrial volume index. DT – deceleration time. KFEI – kinematic filling efficiency index. IS – infarct size. MAR – myocardium at risk.

**Table 4.**
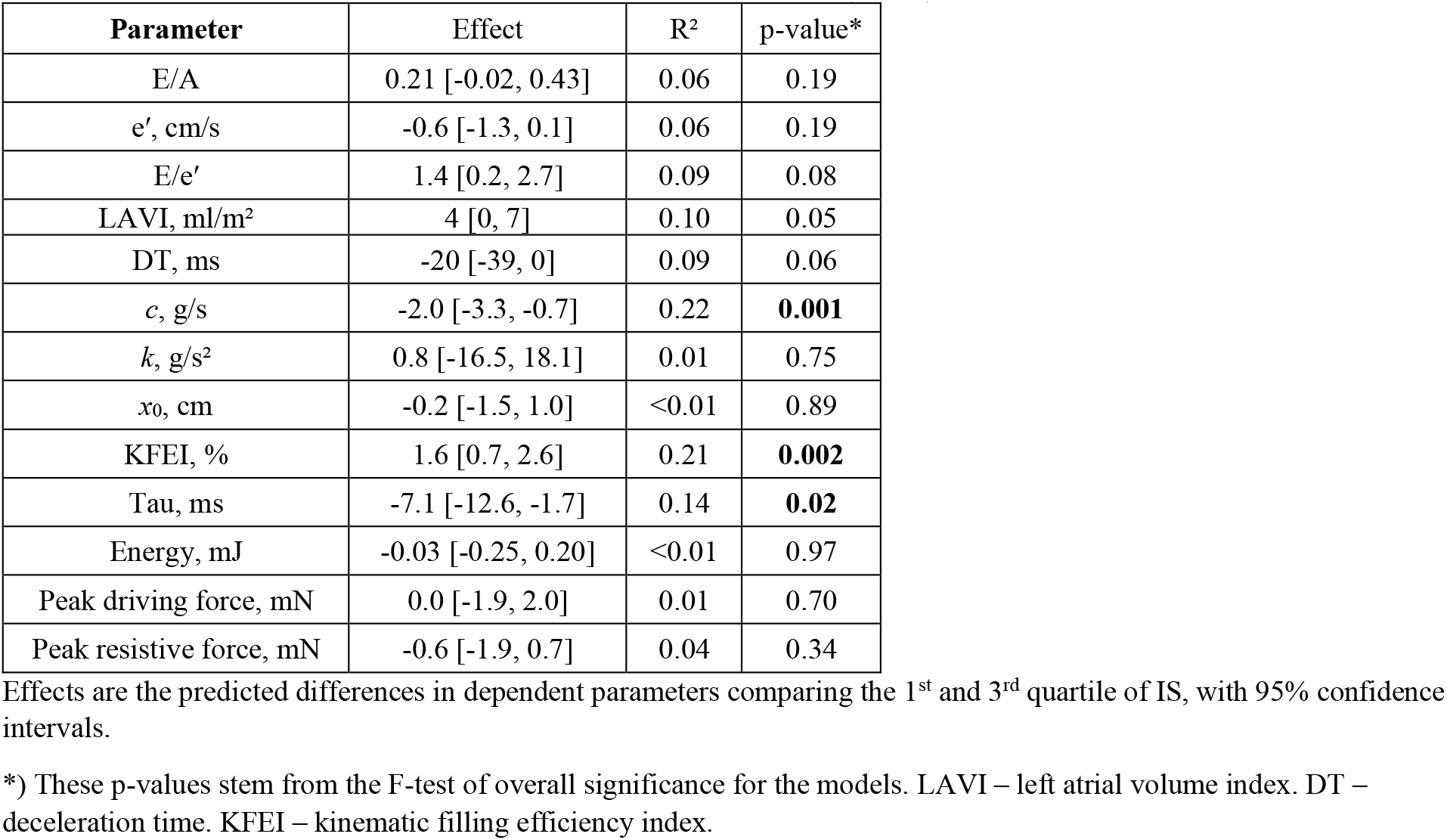
Regression model results, at 6 months follow-up (n=61).

**Figure 1.**
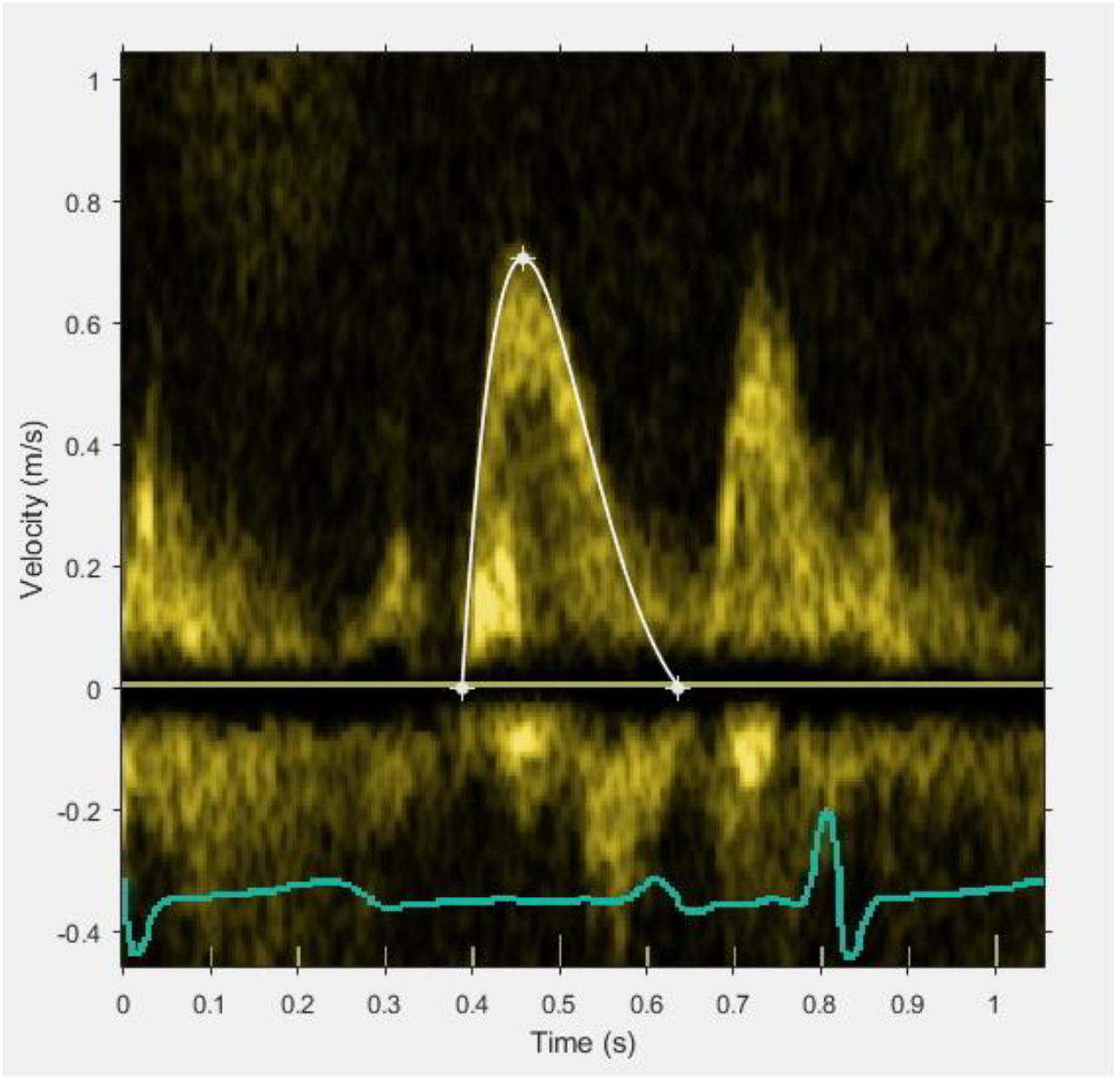
E-wave and PDF fitting example. E-wave as registered by pulsed wave Doppler, superimposed in white is the PDF curve that has been fit to the envelope of the E-wave, and which yielded the constants *x*0=9.0 cm, *c*=20.8 g/s, and *k*=266 g/s^2^.

**Figure 2A.**
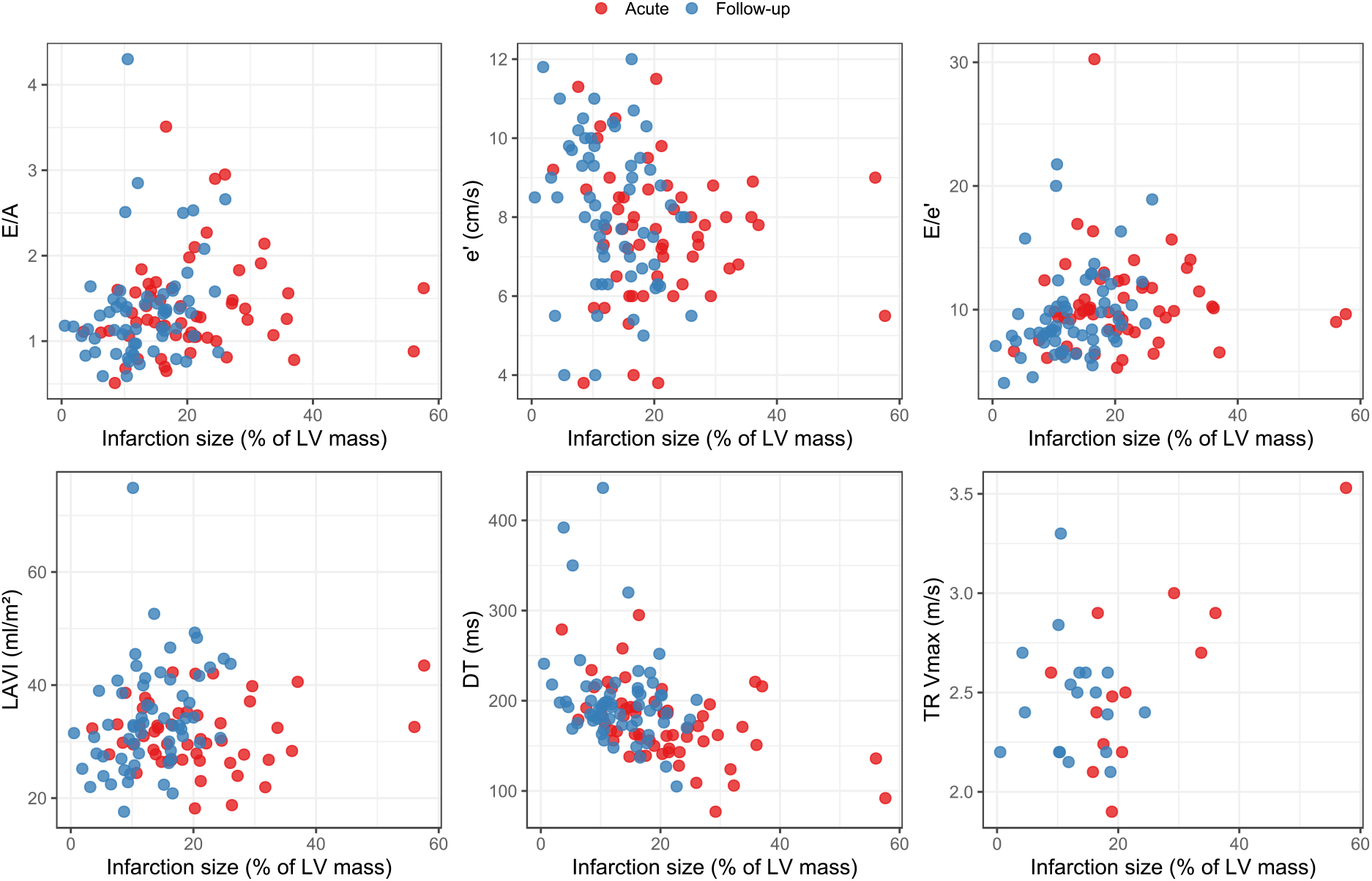
Relationship between infarction size and conventional diastolic parameters acutely (red) and at 6 months follow-up (blue). LAVI – left atrial volume index. DT – deceleration time. TR Vmax – tricuspid regurgitation maximal velocity. LV – left ventricular.

**Figure 2B.**
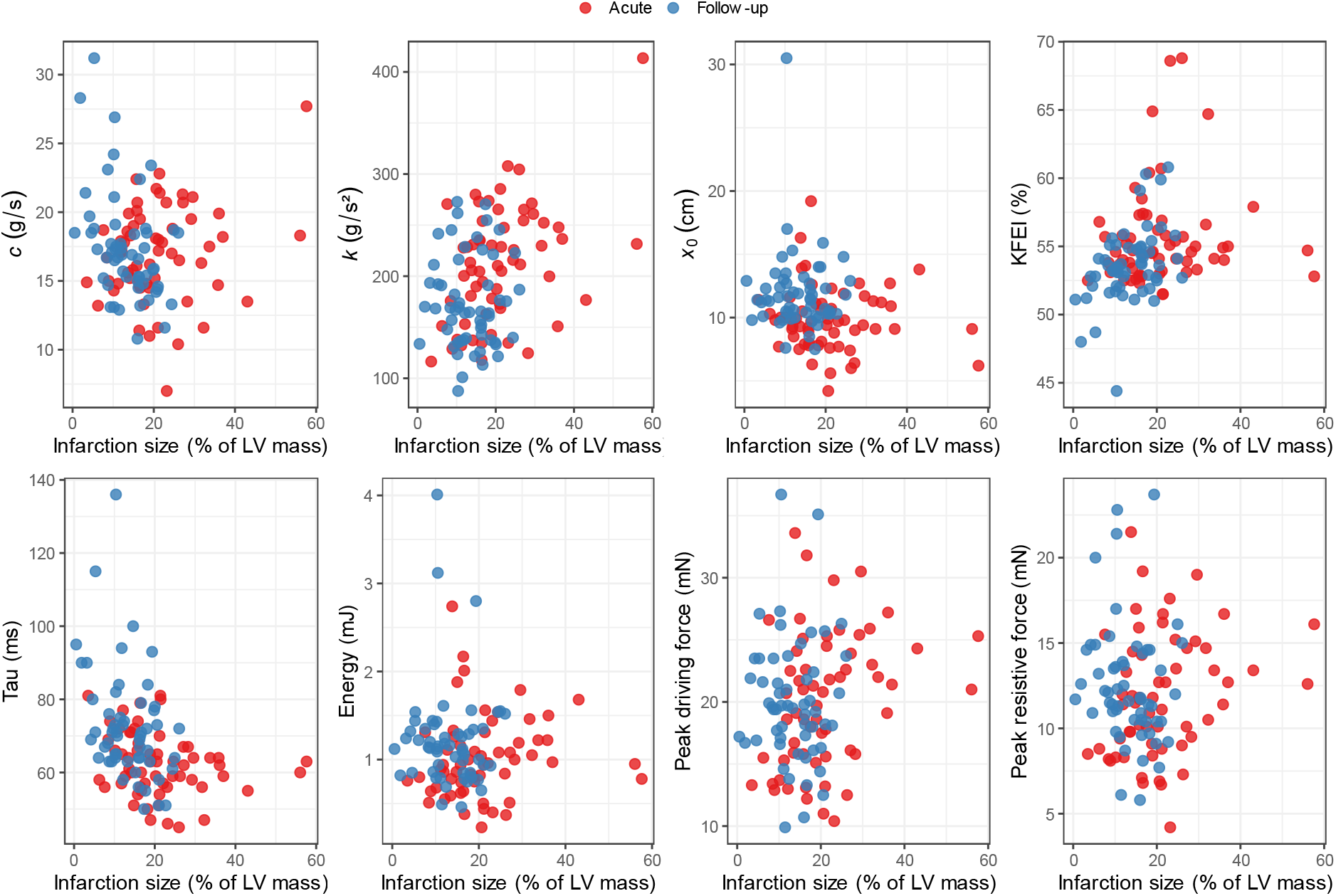
Relationship between infarction size and PDF parameters acutely (red) and at 6 months follow-up (blue). KFEI – kinematic filling efficiency index. LV – left ventricular.

**Figure 3.**
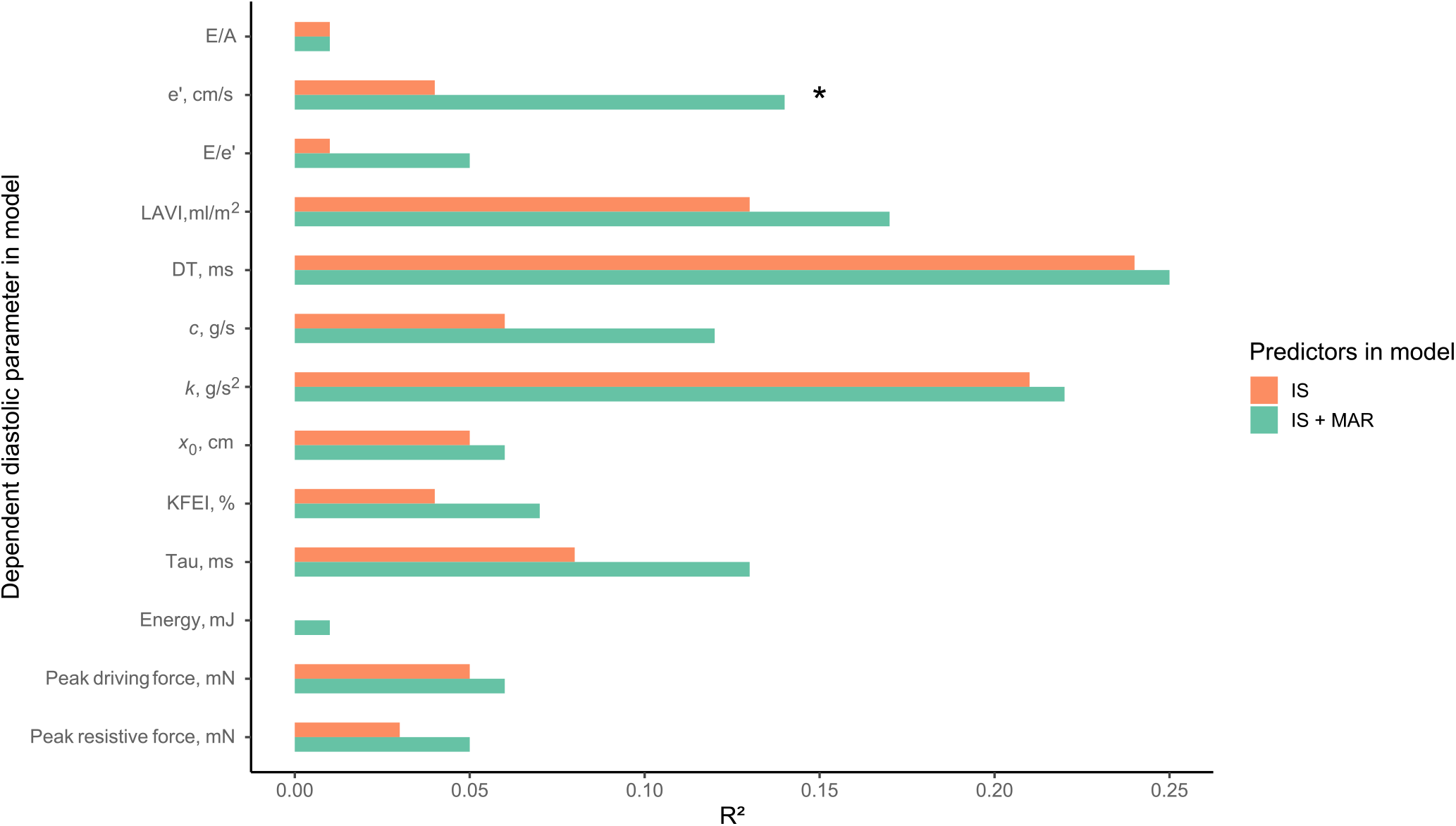
R^2^ for predicting diastolic parameters comparing acute IS to acute IS and MAR. * denotes p<0.05 for the likelihood ratio test. LAVI – left atrial volume index. DT – deceleration time. KFEI – kinematic filling efficiency index.

### Conventional diastolic parameters in the acute setting

Acute IS was associated with left atrial volume index (LAVI) and deceleration time (DT) (R^2^=0.13, p=0.01; R^2^=0.24, p<0.001, respectively), but not with any other conventional parameter. Adding MAR only improved model fit as assessed by a likelihood ratio test in the case of e′ (p=0.02), but the increase in adjusted R^2^ was only 8 percentage points.

### PDF parameters in the acute setting

Acute IS was associated with an increase in myocardial stiffness constant (*k)* (R^2^=0.21, p<0.001), but not with any other PDF parameter. Adding MAR did not improve model fit for any parameter.

### Diastolic parameters at follow-up

Follow-up IS at 6 months was associated with lower *c* (R^2^=0.22, p=0.001) and tau (R^2^=0.14, p=0.02), and higher kinematic filling efficiency index (KFEI, R^2^=0.21, p=0.002), but not with any other parameter.

## Discussion

The main findings of this study are that the association between IS by CMR and various echocardiographic parameters used to assess diastolic function is overall very weak, both in the acute setting and after 6 months. In the acute setting there was a weak association between increasing IS and increasing diastolic stiffness, as manifested by the PDF method’s stiffness parameter *k*, and by decreasing DT using conventional parameters. The edema of the MAR, which in the acute setting could potentially affect myocardial function, did not seem to influence the diastolic parameters in any substantial way, even though the extent of MAR often reached far beyond the extent of the IS. A larger IS at 6 months was associated with a small decrease in viscoelastic energy loss compared to the acute setting, also reflected in the association between IS and Tau and KFEI. For the other diastolic parameters there was no statistically significant association with the final IS observed at follow-up.

The finding of increased myocardial stiffness in the acute setting is in line with early invasive studies of myocardial function during ongoing ischemia^11^, and the overall findings are similar to earlier studies investigating the relationship between IS and various diastolic parameters^6,9^, which found a similarly weak association with IS. Notably, the extent of IS was somewhat greater in the current study (19 [14–27]% or 22±12%), compared to 14 [6–20]% in the study of Barbieri et al^6^ and 15±9% in the study by Chung et al^9^. This suggests that the lack of effect on diastolic dysfunction demonstrated in these studies was not due to a lack of larger MIs. Furthermore, in a study using peak troponin I levels as a proxy for IS^25^, patients with restrictive filling pattern (RFP) and a LVEF >50% had on average only a third of the peak troponin levels as compared with patients with RFP and LVEF <50%, further indicating that the development of diastolic dysfunction after MI is likely to be multifactorial.

By comparison, a recent larger study (n=607) found an association between a semiquantitative assessment of LGE extent and diastolic dysfunction grade^26^. The patient group was heterogeneous, with both ischemic and non-ischemic reasons for exhibiting LGE, and the time elapsed between the disease causing LGE and the time of examination was not taken into consideration, to the extent that it even was known. Given the association between focal myocardial fibrosis and stiffness^27^, it seems inevitable that patients with extensive focal fibrosis would be more prone to have diastolic dysfunction. However, many other factors are likely to influence diastolic dysfunction, including concurrent diseases such as diabetes, hypertension, and heart failure. Notably, the duration of time that a patient has had e.g., hypertension is also highly likely to be of importance, although this information is rarely available. Furthermore, patients can accumulate LGE volume over time, in contrast to the patient population of the present study, which investigated the effect of a first myocardial infarction.

The finding of an association of less viscoelastic energy loss with larger IS at follow-up is novel. However, it seems likely from inspection of the scatter plot that this association was driven by a few individuals with small IS having high *c* at follow-up. This suggests that more data would be of value to better understand the robustness of this relationship.

### Limitations

It is generally agreed that parameters assessing diastolic function are determined by the underlying ability of the heart to fill efficiently as well as the loading conditions at the specific time of registration. Control of loading conditions can only be achieved in a controlled experiment where load is manipulated. We cannot know to what extent loading influenced our results. However, it is not obvious that differences in loading conditions should have obscured a stronger association between IS and diastolic function than the associations found in the current study. Also, it is difficult to ascertain the influence of drug treatment on diastolic function outside the scope of a randomized trial, as treatment indication, dosage, duration, and compliance can vary widely even to the extent that these factors are known. As expected, we did not have a means of controlling for the diastolic function of the included patients prior their enrolment in the RECOND trial but, given their relatively young age, a significant prevalence of diastolic dysfunction seems less likely. We did not analyze the effect of the RECOND trial intervention, partly as the original trial did not show any effect on the pre-specified endpoint, and partly due to the multiplicity testing problem arising in the present study due to the large number of parameters examined.

Another limitation is the sample size, as only 74 patients could be analyzed in the acute setting, and 61 at follow-up. In contrast, a strength of the study was that the range of IS was wide, making the conditions for regression analysis good. Furthermore, our study was limited to a 6-month follow-up. Whether IS could be a factor in long term development of diastolic dysfunction remains to be studied.

## Conclusions

The results of the present study indicate that neither IS nor the myocardial edema in the MAR are major driving forces in the development of diastolic dysfunction after a first anterior MI, and that other factors are likely to be more important. A clinical implication of this would be that when investigating diastolic function with echocardiography after a MI, findings of diastolic dysfunction should not necessarily be interpreted as a sign of a large MI, but, nonetheless, as an indication of increased risk for poor patient outcome.

## Data availability

The data that support the findings of this study are available on reasonable request from the first author.

## References

1. Whalley GA, Gamble GD, Doughty RN. Restrictive diastolic filling predicts death after acute myocardial infarction: systematic review and meta-analysis of prospective studies. Heart Br Card Soc. 2006 Nov;92(11):1588–94.

2. Miller TD, Christian TF, Hopfenspirger MR, Hodge DO, Gersh BJ, Gibbons RJ. Infarct Size After Acute Myocardial Infarction Measured by Quantitative Tomographic 99mTc Sestamibi Imaging Predicts Subsequent Mortality. Circulation. 1995 Aug 1;92(3):334–41.

3. Burns RJ, Gibbons RJ, Yi Q, Roberts RS, Miller TD, Schaer GL, et al. The relationships of left ventricular ejection fraction, end-systolic volume index and infarct size to six-month mortality after hospital discharge following myocardial infarction treated by thrombolysis. J Am Coll Cardiol. 2002 Jan 2;39(1):30–6.

4. Lønborg J, Vejlstrup N, Kelbæk H, Holmvang L, Jørgensen E, Helqvist S, et al. Final infarct size measured by cardiovascular magnetic resonance in patients with ST elevation myocardial infarction predicts long-term clinical outcome: an observational study. Eur Heart J Cardiovasc Imaging. 2013 Apr;14(4):387–95.

5. Johannessen KA, Cerqueira MD, Stratton JR. Influence of myocardial infarction size on radionuclide and Doppler echocardiographic measurements of diastolic function. Am J Cardiol. 1990 Mar 15;65(11):692–7.

6. Barbieri A, Bursi F, Politi L, Rossi L, Fiocchi F, Ligabue G, et al. Echocardiographic diastolic dysfunction and magnetic resonance infarct size in healed myocardial infarction treated with primary angioplasty. Echocardiography. 2008 Jul;25(6):575–83.

7. Laine GA, Allen SJ. Left ventricular myocardial edema. Lymph flow, interstitial fibrosis, and cardiac function. Circ Res. 1991 Jun;68(6):1713–21.

8. Desai KV, Laine GA, Stewart RH, Cox CS Jr, Quick CM, Allen SJ, et al. Mechanics of the left ventricular myocardial interstitium: effects of acute and chronic myocardial edema. Am J Physiol Heart Circ Physiol. 2008 Jun;294(6):H2428–2434.

9. Chung H, Yoon JH, Yoon YW, Park CH, Ko EJ, Kim JY, et al. Different contribution of extent of myocardial injury to left ventricular systolic and diastolic function in early reperfused acute myocardial infarction. Cardiovasc Ultrasound. 2014 Feb 10;12:6.

10. Søholm H, Lønborg J, Andersen MJ, Vejlstrup N, Engstrøm T, Hassager C, et al. Association diastolic function by echo and infarct size by magnetic resonance imaging after STEMI. Scand Cardiovasc J SCJ. 2016 Jun;50(3):172–9.

11. Diamond G, Forrester JS. Effect of Coronary Artery Disease and Acute Myocardial Infarction on Left Ventricular Compliance in Man. Circulation. 1972 Jan 1;45(1):11–9.

12. Kovács SJ Jr, Barzilai B, Pérez JE. Evaluation of diastolic function with Doppler echocardiography: the PDF formalism. Am J Physiol. 1987 Jan;252(1 Pt 2):H178–187.

13. Verouhis D, Sörensson P, Gourine A, Henareh L, Persson J, Saleh N, et al. Effect of remote ischemic conditioning on infarct size in patients with anterior ST-elevation myocardial infarction. Am Heart J. 2016 Nov 1;181:66–73.

14. Heiberg E, Sjögren J, Ugander M, Carlsson M, Engblom H, Arheden H. Design and validation of Segment--freely available software for cardiovascular image analysis. BMC Med Imaging. 2010 Jan 11;10:1.

15. Sörensson P, Heiberg E, Saleh N, Bouvier F, Caidahl K, Tornvall P, et al. Assessment of myocardium at risk with contrast enhanced steady-state free precession cine cardiovascular magnetic resonance compared to single-photon emission computed tomography. J Cardiovasc Magn Reson. 2010;12(1):25.

16. Vermes E, Childs H, Carbone I, Barckow P, Friedrich MG. Auto-threshold quantification of late gadolinium enhancement in patients with acute heart disease. J Magn Reson Imaging JMRI. 2013 Feb;37(2):382–90.

17. Heiberg E, Ugander M, Engblom H, Götberg M, Olivecrona GK, Erlinge D, et al. Automated Quantification of Myocardial Infarction from MR Images by Accounting for Partial Volume Effects: Animal, Phantom, and Human Study. Radiology. 2008 Feb 1;246(2):581–8.

18. Sundqvist MG, Salman K, Tornvall P, Ugander M. Kinematic analysis of diastolic function using the freely available software Echo E-waves - feasibility and reproducibility. BMC Med Imaging. 2016 Oct 27;16(1):60.

19. Zhang W, Chung CS, Riordan MM, Wu Y, Shmuylovich L, Kovacs SJ. The Kinematic Filling Efficiency Index of the Left Ventricle: Contrasting Normal vs. Diabetic Physiology. Ultrasound Med Biol. 2007 Jun;33(6):842–50.

20. Mossahebi S, Kovács SJ. Kinematic Modeling Based Decomposition of Transmitral Flow (Doppler E-Wave) Deceleration Time into Stiffness and Relaxation Components. Cardiovasc Eng Technol. 2014;5(1):25–34.

21. Harrell F. Regression Modeling Strategies: With Applications to Linear Models, Logistic and Ordinal Regression, and Survival Analysis [Internet]. 2nd ed. Springer International Publishing; 2015 [cited 2019 May 3]. (Springer Series in Statistics). Available from: https://www.springer.com/gp/book/9783319194240

22. R Core Team. R: The R Project for Statistical Computing [Internet]. 2019 [cited 2019 May 3]. Available from: https://www.r-project.org/

23. Harrell FE. rms: Regression Modeling Strategies [Internet]. 2019 [cited 2019 May 3]. Available from: https://CRAN.R-project.org/package=rms

24. Wickham H, Chang W, Henry L, Pedersen TL, Takahashi K, Wilke C, et al. ggplot2: Create Elegant Data Visualisations Using the Grammar of Graphics [Internet]. 2019 [cited 2019 May 3]. Available from: https://CRAN.R-project.org/package=ggplot2

25. Prasad SB, Lin A, Kwan C, Sippel J, Younger JF, Hammett C, et al. Determinants of Diastolic Dysfunction Following Myocardial Infarction: Evidence for Causation Beyond Infarct Size. Heart Lung Circ. 2020 Dec;29(12):1815–22.

26. Wang L, Singh H, Mulyala RR, Weber J, Barasch E, Cao JJ. The Association between Left Ventricular Diastolic Dysfunction and Myocardial Scar and Their Collective Impact on All-Cause Mortality. J Am Soc Echocardiogr Off Publ Am Soc Echocardiogr. 2020 Feb;33(2):161–70.

27. Conrad CH, Brooks WW, Hayes JA, Sen S, Robinson KG, Bing OH. Myocardial fibrosis and stiffness with hypertrophy and heart failure in the spontaneously hypertensive rat. Circulation. 1995 Jan 1;91(1):161–70.

