## Supplementary material for "The size of myocardial infarction and peri-infarction edema are not major determinants of diastolic impairment after acute myocardial infarction": RECOND Supplementary material.docx

The PDF method

The PDF method uses the mathematics of damped harmonic motion, a general method for describing the mechanics of a oscillatory movement with decreasing amplitude. The balance of forces of a damped harmonic oscillator are given by

$m\frac{d^{2}x}{dt^{2}}+c\frac{dx}{dt}+kx=0$, (1)

where *m* denotes inertia, *x* displacement from equilibrium, *c* damping, or energy loss, and *k* stiffness. For the purposes of the PDF method, equation 1 is restated as velocity over time as a function of *c*, *k*, and *x*_0_ (*x* at the beginning of motion), and *m* set to 1, expressing the constants as per unit mass. Depending on the relationship between *c* and *k*, three equations are derived. The underdamped case (*c*²-4*k*<0) yields

$v\left( t \right)=\frac{-kx_{0}}{\omega}*\exp\left( \frac{-c}{2}t \right)*sin(\omega t)$, (2)

where ω $=\frac{\sqrt{4k-c^{2}}}{2}$. (3)

The overdamped case (*c*²-4*k*>0) yields

$v\left( t \right)=\frac{-kx_{0}}{\beta}*\exp\left( \frac{-c}{2}t \right)*sinh(\beta t)$ , (4)

where$\beta=\frac{\sqrt{c^{2}-4k}}{2}$ . (5)

The critically damped cases (*c*²-4*k*=0) yields

$v\left( t \right)=-kx_{0}t*\exp\left( \frac{-c}{2}t \right)$ . (6)

The velocity profile of the early mitral inflow (the E-wave) as recorded by PW Doppler constitutes the time-velocity data necessary for curve fitting the appropriate equation. From the curve fitting, the respective values of *c*, *k*, and *x*_0_ which describe the velocity profile of the E-wave are obtained, thus describing the mechanistic properties of energy loss, stiffness, and load during the examined filling phase.
